# Histopathologic Spectrum of Focal Liver Lesions Diagnosed by Ultrasound-Guided Percutaneous Liver Biopsy and Predictors of Hepatocellular Carcinoma: A Single-Center Experience from Sub-Saharan Africa

**DOI:** 10.64898/2026.08.16.26360554

**Authors:** Tamrat Petros Elias, Abate Bane Shewaye, Kaleb Assefa Berhane, Abdu Mohammed, Zebeaman Tibebu, Amanuel Getu Gebreselassie, Abraham Sisay Abie

## Abstract

**Background:** Focal liver lesions (FLLs) encompass a wide spectrum of benign and malignant pathologies, and accurate diagnosis is essential for appropriate management. Although advances in imaging have improved lesion characterization, histopathologic assessment remains the diagnostic gold standard for indeterminate lesions. Data on the histopathologic spectrum and diagnostic utility of ultrasound-guided percutaneous liver biopsy (US-PLB) in sub-Saharan Africa (SSA) are limited. This study aimed to characterize the histopathologic findings of US-PLB performed for FLLs at a tertiary referral center in SSA and to identify factors associated with hepatocellular carcinoma (HCC).

**Methods:** We conducted a retrospective observational study of adult patients (≥18 years) who underwent US-PLB for FLL between January 2021 and December 2024 at Adera Medical and Surgical Center. Patients with indeterminate pathology results, incomplete records, biopsies performed for diffuse liver disease, or lesions classified as LI-RADS 1, 2, or 5 were excluded. Demographic, clinical, laboratory, imaging, histopathologic, and outcome data were extracted from medical records. Descriptive statistics were used to summarize patient characteristics and histopathologic diagnoses. Logistic regression analysis was performed to identify factors associated with HCC.

**Results:** A total of 119 were included in the final analysis. The median age was 56 years (IQR 45– 65), and 59.7% were male. No major biopsy-related complications were reported. HCC was the most common histopathologic diagnosis, accounting for 42.9% of cases, followed by secondary metastatic tumors (15.9%) and regenerative nodules (15.9%). Other diagnoses included chronic hepatitis (8.4%), cholangiocarcinoma (5.9%), and hepatic abscess (3.4%). Hepatitis B virus (HBV) and hepatitis C virus (HCV) infections were present in 14.3% and 12.4% of patients, respectively. On multivariate analysis, HBV infection (AOR 7.85, 95% CI 1.45–42.60; p=0.017), HCV infection (AOR 9.03, 95% CI 1.41–57.76; p=0.020), and larger tumor size (AOR 1.27, 95% CI 1.11–1.46; p<0.01) were significantly associated with HCC.

**Conclusion:** Ultrasound-guided percutaneous liver biopsy demonstrated a favorable safety profile for the evaluation of FLL. HCC was the predominant histopathologic diagnosis, reflecting the substantial burden of primary liver cancer in this setting. Chronic viral hepatitis and larger tumor size were significantly associated with HCC. These findings support the continued role of US-PLB in the diagnostic evaluation of indeterminate focal liver lesions and underscores the importance of viral hepatitis prevention, surveillance, and early detection strategies in sub-Saharan Africa.

## Introduction

Focal liver lesions (FLLs) encompass a broad spectrum of pathologies ranging from benign entities such as cysts, hemangiomas, and focal nodular hyperplasia to primary malignant tumors (HCC and cholangiocarcinoma) and metastatic deposits [1,2]. Accurate characterization of FLLs is essential for directing appropriate management, yet noninvasive imaging like contrast-enhanced ultrasound, multiphasic computed tomography (CT), and magnetic resonance imaging (MRI) may yield indeterminate results, particularly for small lesions (<2 cm) or lesions with atypical enhancement patterns [1–3]. In many clinical scenarios, histopathologic assessment remains the diagnostic reference standard [1,2,4].

Ultrasound-guided percutaneous liver biopsy (US-PLB) provides a minimally invasive means to obtain tissue for definitive diagnosis and for ancillary studies (immunohistochemistry, molecular testing) when imaging is inconclusive or when histologic classification would alter management [1,2,4]. Reported diagnostic yields for image-guided fine-needle aspiration and core-needle biopsy of liver masses are high (sensitivity often >90% and specificity near 100%), while major complication rates are low, typically below 1% in modern series [3,4]. Nevertheless, concerns persist about sampling error, bleeding risk, and rare needle-track tumor seeding; these issues contribute to variation in clinical practice, especially regarding small lesions and patients being evaluated for liver transplantation [1,2,4].

The burden of liver disease in sub-Saharan Africa (SSA) differs from high-income settings. High prevalence of chronic hepatitis B virus (HBV) infection, aflatoxin exposure, limited access to advanced imaging, and frequent late-stage presentation of HCC are common in SSA [5,6]. These region-specific factors could alter the relative frequencies of benign versus malignant FLLs and affect the utility, safety, and diagnostic yield of US-PLB [2,7,8]. However, there is scarcity of published studies describing histopathologic outcomes of US-PLB for FLLs from SSA.

This single-center study aims to characterize the histopathologic findings of ultrasound-guided percutaneous liver biopsies performed for evaluation of focal liver lesions at a tertiary referral center in sub-Saharan Africa. Secondary objectives include documenting indications for biopsy, describing procedure-related safety outcomes, and assessing concordance between pre-biopsy imaging impressions and histopathologic diagnoses. By providing local data on diagnostic yield and complications, we seek to inform context-appropriate biopsy practice and contribute to improved diagnostic pathways for patients with focal hepatic lesions in resource-limited settings.

## Methods

### Study Design and Setting

We conducted a retrospective observational study at the Department of Gastroenterology and Hepatology, Adera Medical and Surgical Center, Addis Ababa, Ethiopia. The study included all consecutive adult patients who underwent US-PLB for FLLs between January 1, 2021, and December 31, 2024.

### Study Population

Adult patients (≥18 years) who underwent US-PLB for one or more focal liver lesions identified on cross-sectional imaging (CT or MRI) were eligible for inclusion. Patients were excluded if histopathologic reports were unavailable or indeterminate, if the biopsy was performed for diffuse liver disease without targeted focal lesion sampling, if clinical records were incomplete, or if lesions had been categorized as Liver Imaging Reporting and Data System (LI-RADS) score 1, 2, or 5 on pre-biopsy imaging.

### Biopsy Procedure and Histopathologic Evaluation

Ultrasound-guided percutaneous liver biopsies were performed by experienced hepatologists according to institutional protocols. Prior to biopsy, all patients underwent routine clinical, laboratory, and imaging assessment, including complete blood count and coagulation profile. Patients with significant ascites, sonographically inaccessible lesions, or coagulation abnormalities (INR >1.5) were not considered suitable for biopsy. Following local anesthesia with 2% lidocaine, tissue samples were obtained under real-time ultrasound guidance using an 18-gauge Tru-cut core biopsy needle. Patients were observed after the procedure for the development of complications. Biopsy specimens were fixed in formalin and processed in the pathology laboratory. Histopathologic evaluation was performed by experienced pathologists using routine hematoxylin and eosin staining.

### Data Collection

Clinical, radiologic, and histopathologic data were extracted from electronic medical records using a standardized data abstraction form between December 22, 2025 and January 23, 2026. Variables collected included demographic characteristics (age and sex), behavioral factors (alcohol consumption, cigarette smoking, and khat chewing), clinical characteristics (comorbidities, symptoms, signs, viral hepatitis status, and presence of cirrhosis), laboratory findings (complete blood count, liver function test, renal function test, and tumor markers), imaging characteristics of focal liver lesions (size, number, and radiologic impression), histopathologic diagnosis, and procedure-related complications. Histopathologic diagnoses were categorized as hepatocellular carcinoma (HCC), metastatic malignancy, cholangiocarcinoma, regenerative nodule, chronic hepatitis, hepatic abscess, or other benign lesions according to the final pathology report.

### Outcome Measures

The primary outcome was the distribution of histopathologic diagnoses among patients undergoing US-PLB for focal liver lesions. Secondary outcomes included identification of factors associated with histopathologically confirmed HCC and assessment of major procedure-related complications including bleeding requiring transfusion or intervention, bile leak, sepsis, or procedure-related death.

### Statistical Analysis

Data were entered, cleaned, and analyzed using SPSS version 25. Continuous variables were summarized using mean ± standard deviation (SD) or median with interquartile range (IQR), depending on data distribution. Categorical variables were presented as frequencies and percentages.

To identify factors associated with HCC, patients with histopathologically confirmed HCC were compared with those with non-HCC diagnoses. Univariable logistic regression analysis was initially performed to estimate crude odds ratios (CORs) and corresponding 95% confidence intervals (CIs). Variables with a p-value <0.20 in univariable analysis, as well as clinically relevant variables identified from the literature, were entered into a multivariable logistic regression model to determine independent predictors of HCC. Adjusted odds ratios (AORs) with 95% CIs were reported. Multicollinearity among predictor variables was assessed using variance inflation factors, and model fit was evaluated using the Hosmer–Lemeshow goodness-of-fit test. Statistical significance was defined as a two-sided p-value <0.05.

### Ethical Considerations

The study was approved by the Institutional Review Board of Adera Medical and Surgical Center and was conducted in accordance with the ethical principles of the Declaration of Helsinki. Written informed consent had been obtained from all patients before the biopsy procedure. Because of the retrospective nature of the study, the requirement for additional informed consent for chart review was waived by the Institutional Review Board. Patient confidentiality was maintained throughout the study by removing names and other identifiers from all data sets.

## Results

During the study period, 133 patients underwent ultrasound-guided percutaneous liver biopsy for the evaluation of focal liver lesions. Of these, 119 patients were included in the final analysis. Nine patients were excluded because of indeterminate histopathologic findings, of which six were due to inadequate tissue sample, and an additional five patients were excluded due to incomplete clinical and laboratory data. No major procedure-related complications were documented during the study period.

### Sociodemographic and Behavioral characteristics

The median age of the study participants was 56 years (IQR: 45–65 years), with ages ranging from 18 to 88 years. Males accounted for 59.7% (n = 71) of the study population, while females comprised 40.3% (n = 48). Regarding behavioral characteristics, alcohol consumption was the most frequently reported exposure, occurring in 10.9% of participants, followed by khat chewing (8.4%) and cigarette smoking (4.2%).

### Clinical characteristics

Overall, 31.9% of participants had at least one comorbid medical condition. Diabetes mellitus was the most common comorbidity, affecting 14.3% of patients, followed by hypertension (10.9%). A family history of liver disease was reported in 3.4% of participants.

Hepatitis B surface antigen (HBsAg) positivity was identified in 14.3% of patients, while 12.4% tested positive for anti-hepatitis C virus antibodies (anti-HCV Ab). Background cirrhosis was present in 26.1% of patients during imaging.

Abdominal pain was the most common presenting symptom, reported by 71.8% of participants, followed by fatigue (45.3%) and weight loss (34.2%). The most frequently observed clinical signs were minimal ascites (13.7%), jaundice (10.3%), and hepatomegaly (7.7%). Notably, 6.0% of patients were asymptomatic, with focal liver lesions detected incidentally during imaging performed for unrelated indications.

### Histopathologic findings

Hepatocellular carcinoma (HCC) was the most common histopathologic diagnosis, accounting for 42.9% of all biopsied lesions. Secondary metastatic tumors and regenerative nodules were the second most frequent diagnoses, each representing 15.9% of cases. Other histopathologic diagnoses included chronic hepatitis (8.4%), cholangiocarcinoma (5.9%), hepatic abscess (3.4%), and focal nodular hyperplasia (1.7%).

**Table 1.** Histopathologic findings of focal liver lesions (n=119)

| Pathology report | Frequency (n) | Percentage (%) |
| --- | --- | --- |
| Hepatocellular carcinoma | 51 | 42.9 |
| Secondary metastasis | 19 | 15.9 |
| Regenerative nodules | 19 | 15.9 |
| Chronic hepatitis | 10 | 8.4 |
| Cholangiocarcinoma | 7 | 5.9 |
| Hepatic abscess | 4 | 3.4 |
| Focal nodular hyperplasia | 2 | 1.7 |
| Hepatic adenoma | 2 | 1.7 |
| Others | 5 | 4.2 |

### Factors associated with Histopathologic Diagnosis of HCC

On univariable logistic regression analysis, HBsAg positivity (COR 1.78, 95% CI: 1.20–2.64; p = 0.020), anti-HCV antibody positivity (COR 1.91, 95% CI: 1.29–2.82; p = 0.013), background cirrhosis (COR 1.54, 95% CI: 1.03–2.31; p = 0.040), and larger tumor size (COR 1.21, 95% CI: 1.08–1.35; p < 0.001) were significantly associated with a histopathologic diagnosis of HCC.

Variables meeting the predefined criteria for inclusion in the multivariable model were subsequently entered into a multivariable logistic regression analysis. After adjustment for potential confounders, HBsAg positivity (AOR 7.85, 95% CI: 1.45–42.60; p = 0.017), anti-HCV antibody positivity (AOR 9.03, 95% CI: 1.41–57.76; p = 0.020), and tumor size (AOR 1.27, 95% CI: 1.11–1.46; p < 0.001) remained independently associated with HCC. Background cirrhosis was no longer statistically significant after adjustment for other covariates.

**Table 2.** Factors associated with HCC diagnosis.

| Character | COR (95% CI) | P-Value | AOR (95% CI) | P-Value |
| --- | --- | --- | --- | --- |
| HBsAg Positive | 1.78 (1.20–2.64) | 0.02 | 7.85(1.45-42.60) | 0.01 |
| Anti-HCV Ab Positive | 1.91 (1.29–2.82) | 0.013 | 9.03(1.41-57.76) | 0.02 |
| Background cirrhosis | 1.54 (1.03–2.31) | 0.04 | 1.30(0.35-4.84) | 0.69 |
| Tumor size | 1.21 (1.08–1.35) | <0.01 | 1.27(1.11-1.46) | <0.01 |
| Abbreviations: HBsAg – Hepatitis B Surface antigen; HCV Ab – Hepatitis C virus antibody |  |  |  |  |

## Discussion

In this retrospective study of 119 patients who underwent ultrasound-guided percutaneous liver biopsy for focal liver lesions, hepatocellular carcinoma was the most common histopathologic diagnosis, accounting for nearly half of all lesions, followed by metastatic tumors and regenerative nodules. Chronic viral hepatitis was prevalent in the study population, with hepatitis B virus and hepatitis C virus infection identified in 14.3% and 12.4% of patients, respectively, while approximately one-quarter of patients had underlying cirrhosis. Abdominal pain was the predominant presenting symptom, although a small proportion of lesions were detected incidentally. Importantly, no major procedure-related complications were observed. Hepatitis B virus infection, hepatitis C virus infection, and increasing tumor size were independently associated with a histopathologic diagnosis of hepatocellular carcinoma.

The predominance of HCC observed in this study is consistent with the epidemiology of liver cancer in sub-Saharan Africa, where HCC remains the most common primary hepatic malignancy and contributes substantially to cancer-related mortality [9]. Chronic viral hepatitis continues to be the leading etiologic factor for HCC in the region, while delayed presentation and limited surveillance programs frequently result in diagnosis at advanced stages [10]. Recent reviews have shown that HBV related HCC remains highly prevalent across sub-Saharan Africa and that the majority of patients present with symptomatic disease, often precluding curative treatment options [11]. The high proportion of HCC in our cohort likely reflects both the underlying burden of chronic viral hepatitis and the referral nature of our institution, where patients with suspicious focal liver lesions are commonly evaluated.

Metastatic tumors represented the second most common histopathologic diagnosis in our study. This finding is expected as liver is a frequent site of hematogenous metastasis because of its dual blood supply from the portal vein and hepatic artery. Although imaging modalities such as multiphasic CT and MRI have improved characterization of liver lesions, distinguishing primary liver malignancies from metastatic disease remains challenging in many cases, particularly when lesions demonstrate atypical imaging characteristics. Histopathologic confirmation therefore continues to play a critical role in establishing definitive diagnoses and guiding treatment decisions. Previous studies have similarly reported that metastatic lesions constitute a substantial proportion of biopsied focal liver lesions, particularly in tertiary referral centers [12,13].

An important finding of the present study was the strong association between HBV infection and HCC. After adjustment for potential confounders, patients with positive HBsAg had nearly eight-fold higher odds of HCC compared with HBV-negative individuals. This observation is biologically plausible and supported by extensive epidemiologic evidence demonstrating that chronic HBV infection is one of the strongest risk factors for HCC [14,15]. Unlike many other causes of chronic liver disease, HBV may promote HCC both indirectly through cirrhosis and directly through viral DNA integration into the host genome, resulting in genetic instability and malignant transformation [16,17]. HBV-associated HCC may also occur in the absence of established cirrhosis, a phenomenon frequently reported in African populations [18]. The magnitude of association observed in our study underscores the continuing need for HBV screening, antiviral treatment, and HCC surveillance programs in resource-limited settings.

Similarly, HCV infection was independently associated with HCC, with affected patients demonstrating approximately nine-fold increased odds of malignancy. Chronic HCV infection contributes to progressive hepatic inflammation, fibrosis, and cirrhosis, ultimately increasing the risk of HCC development [19]. Although HCV prevalence in SSA is generally lower than HBV prevalence, HCV remains an important contributor to the regional burden of liver cancer [20, 21]. The association observed in our study is consistent with previous investigations demonstrating that chronic HCV infection significantly increases the likelihood of HCC among patients with focal liver lesions.

Tumor size emerged as another independent predictor of HCC. Each incremental increase in lesion size was associated with higher odds of a malignant diagnosis. This finding is clinically relevant because larger lesions are more likely to represent aggressive biological behavior and advanced disease. While small hepatic lesions can represent a broad spectrum of benign and malignant conditions, increasing lesion size has consistently been associated with a greater probability of malignancy in imaging and pathology studies [22]. In many low-resource settings, delays in access to diagnostic imaging and specialist care may contribute to presentation with larger tumors, thereby increasing the likelihood of HCC diagnosis at the time of biopsy.

Interestingly, cirrhosis was associated with HCC in univariable analysis but lost statistical significance after multivariable adjustment. This finding may reflect the strong correlation between cirrhosis and chronic viral hepatitis within the study population. Because HBV and HCV are major causes of both cirrhosis and HCC, adjustment for viral hepatitis status may have attenuated the independent effect of cirrhosis. Alternatively, the relatively modest sample size may have limited statistical power to detect independent associations among closely related variables. Nevertheless, the well-established role of cirrhosis as a major precursor of HCC remains widely recognized. It is worth noting, however, that recent studies have suggested that the absolute risk of HCC among patients with compensated cirrhosis may be lower than previously reported, particularly in the era of effective antiviral therapy, improved surveillance, and better management of underlying liver diseases. These findings indicate that while cirrhosis remains an important risk factor, the magnitude of its contribution to HCC development may vary according to etiology, disease stage, and access to contemporary treatment strategies [23,24]. Therefore, the lack of an independent association in our multivariable model should be interpreted cautiously and does not diminish the established biological relationship between cirrhosis and HCC.

No major complications were documented following ultrasound-guided percutaneous liver biopsy. This finding is consistent with contemporary evidence demonstrating that image-guided liver biopsy is a safe procedure when performed by experienced operators following appropriate patient selection and pre-procedural assessment. A systematic review and meta-analysis involving more than 12,000 patients reported extremely low rates of major complications after ultrasound-guided liver biopsy, while other reviews have estimated major complication rates of less than 1% [25,26]. The absence of major complications in our study further supports the continued use of ultrasound-guided biopsy for the evaluation of focal liver lesions when histopathologic confirmation is clinically indicated.

The findings of this study should be interpreted in light of several limitations. First, the retrospective design may have introduced information bias due to incomplete documentation of clinical and behavioral variables. Second, the study was conducted at a single tertiary referral center, which may limit the generalizability of the findings to other settings. Third, patients with indeterminate pathology results and incomplete records were excluded, potentially introducing selection bias. Finally, the relatively small sample size may have reduced the ability to detect weaker associations between clinical variables and HCC.

Despite these limitations, this study provides important data on the histopathologic spectrum of focal liver lesions in a sub-Saharan African setting and identifies key factors associated with HCC. The findings highlight the substantial burden of HCC among patients undergoing liver biopsy and emphasize the critical role of chronic HBV and HCV infections in hepatocarcinogenesis. Strengthening viral hepatitis prevention, early diagnosis, antiviral treatment, and HCC surveillance programs may contribute substantially to reducing the burden of liver cancer in the region.

## Conclusion

In this single-center retrospective study, hepatocellular carcinoma was the most common histopathologic diagnosis among patients undergoing ultrasound-guided percutaneous liver biopsy for focal liver lesions, followed by metastatic tumors and regenerative nodules. Chronic hepatitis B virus infection, hepatitis C virus infection, and larger tumor size were independently associated with a histopathologic diagnosis of HCC. Ultrasound-guided percutaneous liver biopsy demonstrated an excellent safety profile, with no major procedure-related complications observed during the study period.

These findings highlight the substantial burden of viral hepatitis-associated liver cancer in sub-Saharan Africa and underscore the importance of strengthening HBV and HCV screening, surveillance, and treatment programs. Furthermore, our results support the continued use of ultrasound-guided percutaneous liver biopsy as a safe and valuable diagnostic tool for the evaluation of indeterminate focal liver lesions, particularly in resource-limited settings where accurate histopathologic diagnosis remains essential for guiding clinical management.

## Data Availability

I confirm that all relevant data will be made available in accordance with the journal Data Availability policy

## Acknowledgement

We would like to thank the management and staffs of Adera medical and surgical center for their support throughout the study.

## Reference

1. 1. Frenette C, Mendiratta-Lala M, Salgia R, Wong RJ, Sauer BG, Pillai A. ACG clinical guideline: focal liver lesions. Official journal of the American College of Gastroenterology| ACG. 2024 Jul 1;119(7):1235–71.

2. Tapper EB, Lok AS. Use of liver imaging and biopsy in clinical practice. N Engl J Med. 2017;377(8):756–768. doi:10.1056/NEJMra1610570.

3. Pöschel T, Blank V, Schlosser T, Lingscheidt T, Böhlig A, Wiegand J, Karlas T. Ultrasound-guided percutaneous biopsy for focal liver lesions: adverse events and diagnostic yield in a single-centre analysis. PLoS One. 2020;15(3):e0230129. doi:10.1371/journal.pone.0230129.

4. Boyum JH, Atwell TD, Schmit GD, Poterucha JJ, Schleck CD, Harmsen WS, et al. Incidence and risk factors for adverse events related to image-guided liver biopsy. Mayo Clin Proc. 2016;91(3):329–335. doi:10.1016/j.mayocp.2015.12.012.

5. Spearman CW. The burden of chronic liver disease in West Africa: a time for action. The Lancet Global Health. 2023 Sep 1;11(9):e1319–20.

6. Wu XN, Xue F, Zhang N, Zhang W, Hou JJ, Lv Y, Xiang JX, Zhang XF. Global burden of liver cirrhosis and other chronic liver diseases caused by specific etiologies from 1990 to 2019. BMC Public Health. 2024 Feb 3;24(1):363.

7. Russo MW, Shrestha R. Evaluation of Focal Liver Masses. Gastrointestinal and Liver Secrets E-Book: Gastrointestinal and Liver Secrets E-Book. 2023 Dec 16:185.

8. Khalifa A, Sasso R, Rockey DC. Role of liver biopsy in assessment of radiologically identified liver masses. Digestive diseases and sciences. 2022 Jan;67(1):337–43.

9. Younossi ZM, Wong G, Anstee QM, Henry L. The global burden of liver disease. Clinical Gastroenterology and Hepatology. 2023 Jul 1;21(8):1978–91.

10. Mbaga DS, Kenmoe S, Kengne-Ndé C, Ebogo-Belobo JT, Mahamat G, Foe-Essomba JR, Amougou-Atsama M, Tchatchouang S, Nyebe I, Feudjio AF, Kame-Ngasse GI. Hepatitis B, C and D virus infections and risk of hepatocellular carcinoma in Africa: A meta-analysis including sensitivity analyses for studies comparable for confounders. PLoS One. 2022 Jan 21;17(1):e0262903.

11. Sobnach S, Kotze U, Spearman CW, Sonderup M, Nashidengo PR, Ede C, Keli E, Chihaka O, Zerbini LF, Li YJ, Gandhi K. The management and outcomes of hepatocellular carcinoma in sub-Saharan Africa: a systematic review. HPB. 2024 Jan 1;26(1):21–33.

12. Hoffmann P, Cyrany J, Kopecky J, Hoffmannova M, Ryska P, Hulek M, Dvorak P. Percutaneous CT-guided Biopsy of Focal Liver Lesions--Longterm Experience with more than 300 Procedures. Journal of Gastrointestinal & Liver Diseases. 2023 Jun 1;32(2).

13. Ilie M, Rusu M, Rosianu C, Neagu TP, Motofei IG, Bratu OG, Socea B, Stanescu AM, Gherghiceanu F, Pantea Stoian A. Ultrasound-guided biopsy in focal liver lesions. Arch Balk Med Union. 2018 Sep;53(3):364–8.

14. Duberg AS, Lybeck C, Fält A, Montgomery S, Aleman S. Chronic hepatitis B virus infection and the risk of hepatocellular carcinoma by age and country of origin in people living in Sweden: a national register study. Hepatology communications. 2022 Sep;6(9):2418–30.

15. Lin CL, Kao JH. Development of hepatocellular carcinoma in treated and untreated patients with chronic hepatitis B virus infection. Clinical and Molecular Hepatology. 2023 Feb 15;29(3):605.

16. Rizzo GE, Cabibbo G, Craxi A. Hepatitis B virus-associated hepatocellular carcinoma. Viruses. 2022 May 7;14(5):986.

17. Tuemen D, Heumann P, Guelow K, Demirci CN, Cosma LS, Mueller M, Kandulski A. Pathogenesis and current treatment strategies of hepatocellular carcinoma. Biomedicines. 2022 Dec 9;10(12):3202.

18. Matthews PC, Kramvis A. Hepatitis B virus (HBV) and hepatocellular carcinoma (HCC) in sub-Saharan Africa: no room for complacency. Hepatoma Research. 2022 Mar 26;8(3):14.

19. Fiehn F, Beisel C, Binder M. Hepatitis C virus and hepatocellular carcinoma: carcinogenesis in the era of direct-acting antivirals. Current Opinion in Virology. 2024 Aug 1;67:101423.

20. Kassa GM, Walker JG, Alamneh TS, Tamiru MT, Bivegete S, Adane A, Amogne W, Dillon JF, Vickerman P, Dagne E, Yesuf EA. Prevalence, trends, and distribution of hepatitis C virus among the general population in sub-Saharan Africa: A systematic review and meta-analysis. Liver International. 2024 Dec;44(12):3238–49.

21. Mbaga DS, Kenmoe S, Kengne-Ndé C, Ebogo-Belobo JT, Mahamat G, Foe-Essomba JR, Amougou-Atsama M, Tchatchouang S, Nyebe I, Feudjio AF, Kame-Ngasse GI. Hepatitis B, C and D virus infections and risk of hepatocellular carcinoma in Africa: A meta-analysis including sensitivity analyses for studies comparable for confounders. PLoS One. 2022 Jan 21;17(1):e0262903.

22. Mbaga DS, Kenmoe S, Kengne-Ndé C, Ebogo-Belobo JT, Mahamat G, Foe-Essomba JR, Amougou-Atsama M, Tchatchouang S, Nyebe I, Feudjio AF, Kame-Ngasse GI. Hepatitis B, C and D virus infections and risk of hepatocellular carcinoma in Africa: A meta-analysis including sensitivity analyses for studies comparable for confounders. PLoS One. 2022 Jan 21;17(1):e0262903.

23. Kanwal F, Khaderi S, Singal AG, Marrero JA, Loo N, Asrani SK, Amos CI, Thrift AP, Gu X, Luster M, Al-Sarraj A. Risk factors for HCC in contemporary cohorts of patients with cirrhosis. Hepatology. 2023 Mar 1;77(3):997–1005.

24. Zhang Y, Liu X, Li S, Lin C, Ye Q, Wang Y, Wu J, Zhang Y, Gao H, Li T, Qu Y. Risk of HCC decreases in HBV-related patients with cirrhosis acquired recompensation: A retrospective study based on Baveno VII criteria. Hepatology communications. 2024 Jan 1;8(1):e0355.

25. Tian G, Kong D, Jiang TA, Li L. Complications after percutaneous ultrasound-guided liver biopsy: a systematic review and meta-analysis of a population of more than 12,000 patients from 51 cohort studies. Journal of Ultrasound in Medicine. 2020 Jul;39(7):1355–65.

26. Chai WL, Lu DL, Sun ZX, Cheng C, Deng Z, Jin XY, Zhang TL, Gao Q, Pan YW, Zhao QY, Jiang TA. Major complications after ultrasound-guided liver biopsy: An annual audit of a Chinese tertiary-care teaching hospital. World Journal of Gastrointestinal Surgery. 2023 Jul 27;15(7):1388.

